# Performance in Fundamentals of Laparoscopic Surgery (FLS): Does it Reflect Global Rating Scales in Objective Structured Assessment of Technical Skills (OSATS) in Porcine Laparoscopic Surgery?

**DOI:** 10.1101/2022.03.31.22273188

**Authors:** Ho Yee Tiong, Wei Zheng So, Jeremy Yuen-Chun Teoh, Shuji Isotani, Gang Zhu, Teng Aik Ong, Eddie Shu-Yin Chan, Peggy Sau-Kwan Chu, Kittinut Kijvikai, Ming Liu, Bannakji Lojanapiwat, Michael Wong, Anthony Chi-Fai Ng

## Abstract

**Background:** To correlate the utility of Fundamentals of Laparoscopic Surgery (FLS) manual skills program with the Objective Structured Assessment of Technical Skills (OSATS) global rating scale in evaluating operative performance.

**Materials and Methods:** The Asian Urological Surgery Training and Educational Group (AUSTEG) Laparoscopic Upper Tract Surgery Course (LUTSC) implemented and validated the FLS program for its usage in laparoscopic surgical training. Delegates’ basic laparoscopic skills were assessed using three different training models (Peg Transfer, Precision Cutting and Intra-corporeal Suturing). They also performed live porcine laparoscopic surgery at the same workshop. Live surgery skills were assessed by blinded faculty using the OSATS rating scale.

**Results:** From 2016 to 2019, a total of 81 certified urologists participated in the course, with a median of 5 years’ experience post residency. Although differences in task timings did not reach statistical significance, those with more surgical experience were visibly faster at completing the peg transfer and intra-corporeal suturing FLS tasks. However, they took longer to complete the precision cutting task than participants with less experience. Overall OSATS scores correlated weakly with all three FLS tasks (Peg Transfer Time: R = -0.331, R2 = 0.110; Precision Cutting Time: R = - 0.240, R2 = 0.058; Suturing with Intra-corporeal Knot Time: R = -0.451, R2 = 0.203).

**Conclusion:** FLS task parameters did not correlate strongly with OSATS globing rating scale performance. Although the FLS task models demonstrated strong validity, it is important to assimilate the inconsistencies when benchmarking technical proficiency against real-life operative competence, as evaluated by FLS and OSATS respectively.

## INTRODUCTION

Initially developed by The Society of American Gastrointestinal and Endoscopic Surgeons (SAGES) in 1997 and subsequently validated by the American College of Surgeons, the Fundamentals of Laparoscopic Surgery (FLS) program^1^ is an educational framework that seeks to inculcate the foundations of technical proficiency specific to laparoscopic surgery, holistically encompassing multiple aspects of learning – didactic teachings, manual skills tasks and an evaluation component. Since its advent into the surgical community, the FLS program has been rigorously implemented and endorsed across various subspecialties that demonstrate a wide range of laparoscopic surgery skillsets^2-4^, reaffirming its continued validity in contemporary operative techniques.

The manual skills component is intended to assess the extent of one ’s technical abilities whilst performing basic laparoscopic surgical maneuvers. Pioneered by Dr. Gerald Fried in the McGill Inanimate System for Training and Evaluation of Laparoscopic Skills (MISTELS)^5^, it has since been tailored to the specifics of the FLS system and deemed as a mandatory board eligibility pre-requisite for all general surgery residents in the United States. Evaluation parameters are determined by the duration that the candidate takes to fulfil the stipulated tasks, with a specified maximum time limit for completion.

In the same vein, the Objective Structured Assessment of Technical Skills (OSATS)^6, 7^ is another widely incorporated objective tool that aims to evaluate model-based or live surgery performance within the operating theatre. It was first established by Martin et al. in the University of Toronto back in 1997 and has since been labelled as a reliable and validated tool for technical skill assessment in surgical expertise. Notably, it provides a 7-item global rating checklist graded on a 5-point Likert scale that is universally applicable across all surgical disciplines and has been demonstrated to adequately reflect the level of one’s technical competency in surgical skills^8, 9^.

The transition of performing surgery on inanimate models to the live operating field has always been regarded as a challenging process with a steep learning curve to master, with surfacing concerns such as the replicability of skillsets and an unpredictable operating environment experienced in live surgery. Although bench simulation training has been shown to correlate with real-life surgical performance^10-13^ in several noteworthy studies, some inconsistencies in current literature have been observed that report otherwise^14, 15^. Henceforth, it remains relatively unclear as to whether surgical simulator models can accurately determine actual surgical finesse in the operating room.

As a surrogate measure of laparoscopic proficiency on simulator models, the Asian Urological Surgery Training and Education Group (AUSTEG) sought to comparatively assess the reliability of FLS manual task parameters and its ability to predict the performance of OSATS scores measured on the global rating scale. The group has since validated and published evidence on several task models for effective training within the urological context^16, 17^. To analyze our hypothesis, we organized and conducted an annual Laparoscopic Upper Tract Surgery Course that was aimed at assessing attendees’ basic laparoscopic skills in urology. The program has consistently garnered overwhelming interest and positive feedback since its inception in 2016.

## METHODS

AUSTEG is a non-profit training organization, founded in November 2015 with a faculty comprising of expert urologists from more than 10 Asian countries. A plethora of training courses are conducted every year with the goal of priming budding urologists in Asia with relevant knowledge and skills on surgical techniques, focusing on key categories such as Laparoscopic Upper Tract Surgery, Lower Urinary Tract Endourology, Endoscopic Stone Management and Pelvic Laparoscopic Surgery.

### AUSTEG Laparoscopic Upper Tract Surgery Course

From March 2016 to 2019, Urological residents and delegates were invited to participate in the annual Laparoscopic Upper Tract Surgery course that was first held at the Olympus China Medical Training and Education Centre (C-TEC) in Guangzhou, China. During the two-day course, they had to perform three FLS simulator tasks – Peg Transfer, Precision Cutting and Suturing with Intra-corporeal knot. Subsequently, they were tasked to complete three laparoscopic live porcine surgery urological procedures – uretero-ureterotomy, ureteric re-implantation and total nephrectomy.

Prior to the session, participants were provided with a weblink showcasing the demonstration of the FLS manual tasks, as well as standardized instructional step-by-step video techniques of the three procedures respectively. A validation assessment of the FLS task models was also carried out, whereby participants were asked to fill in both pre- and post-task questionnaires to determine the face and content validity of each simulation task.

For each FLS task, participants executed each station under timed conditions and the duration taken to fulfil the requirements of each task was recorded. Afterwards, participants were paired out for the three live porcine surgical procedures. Pertaining to the live surgical performance evaluation of these delegates, the evaluating faculty was blinded to the delegates’ previous background and experience, as well as their performance parameters attained in the three FLS manual tasks.

The pre-task questionnaire, post-task questionnaire and the OSATS global rating scales were all issued and assessed by the AUSTEG faculty, who are academic urologists actively involved in and representing accredited urology residency programs from different Asian countries. They are therefore familiar with competency-based assessment and equipped with the relevant expertise to objectively assess the participants’ performance during live porcine laparoscopic surgery.

Participants were stratified based on the number of real-life laparoscopic nephrectomies performed prior to attending the course as an indicator of surgical experience. A cut-off of 10 laparoscopic nephrectomies was adopted in accordance with a previously published paper that depicted consistency of intra-operative and post-operative parameters after participation in the tenth laparoscopic case onwards^18^.

### Statistical Analysis

χ^2^ tests (or Fisher’s Exact Test, wherever applicable) and Independent T-test were used to compare between categorical and continuous variables respectively. All *p*-values were two-tailed, and a *p-*value of <0.05 was taken to be statistically significant. Categorical variables are reported as number (percent) while continuous variables are reported as mean (standard deviation; SD). Relationships between individual FLS task timings and the mean overall OSATS score achieved by participants were investigated using linear correlation, utilizing the coefficient of determination (R; Pearson correlation coefficient) as a measure of the goodness of fit. Absolute R values of > 0.8 were considered as strong correlations between the two variables.

Multivariate regression analysis was carried out to examine the association between overall OSATS scores and predictor variables (Peg Transfer Timing, Precision Cutting Timing, Suturing with Intra-corporeal Knot Timing), and the coefficient of determination (R^2^) were used as a measure of the extent of correlation between FLS task parameters and overall OSATS scores. All statistical analyses were performed using IBM SPSS Version 26.0.

## RESULTS

### Baseline Characteristics

From 2016 to 2019, a total of 128 certified urologists participated in the course. 47 participants were excluded from analysis due to incomplete responses. There were 81 urologists (Male = 74, Female = 7) with a median of 5 years’ experience post-residency. Baseline characteristics are depicted in Table 1. Pre and post-task questionnaires were administered to evaluate the utility of the three simulator models, with responses recorded on a 5-point Likert Scale. All demonstrated excellent acceptability, face and content validity (Figure 1).

**Table 1:** Baseline Characteristics of Participating Urologists from 2016 to 2019

|  | **≤ 10 Lap**<br>**Nephrectomies** | **> 10 Lap**<br>**Nephrectomies** | **p-**<br>**value** |
| --- | --- | --- | --- |
|  | (n = 54) | (n = 27) |  |
| Gender [n (%)] |  |  | >0.05 |
| Male | 49 (90.7) | 25 (92.6) |  |
| Female | 5 (9.3) | 2 (7.4) |  |
| Designation [n (%)] |  |  | 0.914 |
| Resident | 2 (3.7) | 0 (0) |  |
| Consultant | 30 (55.6) | 16 (59.3) |  |
| Senior<br>Consultant | 22 (40.7) | 11 (40.7) |  |
| Have Performed at<br>Least 1<br>Laparoscopic<br>Nephrectomy in the<br>Past Six Months [n<br>(%)] |  |  | 0.547 |
| Yes | 51 (94.4) | 27 (100) |  |
| No | 3 (5.6) | 0 (0) |  |

**Figure 1:**
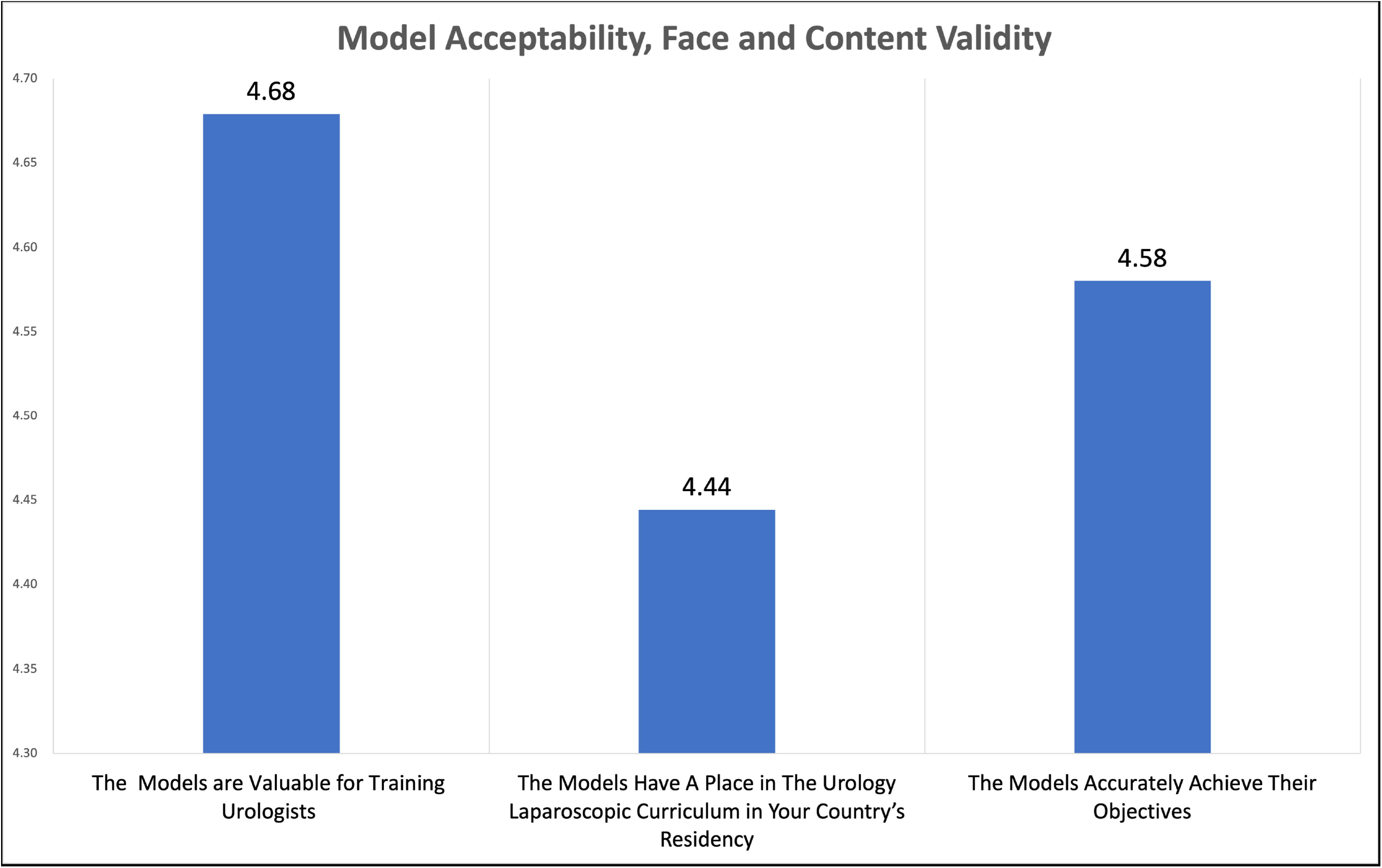
Questionnaire Demonstrating Validity of FLS Task Models

### FLS and OSATS Parameters

54 delegates performed ≤10 real-life nephrectomies prior to attending the course, while the remaining 27 performed >10 real-life nephrectomies. Performance timings for all three FLS tasks did not differ significantly between participants of varying experience levels (Table 2). Those with more surgical experience demonstrated shorter timings for the peg transfer (148.8 ± 52.4 vs. 138.4 ± 56.7 seconds, p = 0.200) and intra-corporeal suturing (280.7 ± 169.8 vs. 247.1 ± 152.4 seconds, p = 0.296) FLS tasks. However, they took longer to complete the pattern cut task (252.4 ± 144.7 vs. 232.3 ± 112.2 seconds, p = 0.767) than participants with less experience.

**Table 2:** FLS Task Parameters

|  | **≤ 10 Lap**<br>**Nephrectomies** | **> 10 Lap**<br>**Nephrectomies** | **p-value** |
| --- | --- | --- | --- |
|  | (n = 54) | (n = 27) |  |
| Mean Peg Transfer<br>Timing, seconds [mean<br>(SD)] | 148.8 (52.4) | 138.4 (56.7) | 0.200 |
| Mean Precision Cutting<br>Time, seconds [mean<br>(SD)] | 232.3 (112.2) | 252.4 (144.7) | 0.767 |
| Intra-corporeal<br>Suturing Time,<br>seconds [mean (SD)] | 280.7 (169.8) | 247.1 (152.4) | 0.296 |

Likewise, no differences were seen between the two groups across the individual components of the OSATS score, as well as the overall score (Table 3).

**Table 3:** OSATS Evaluation Parameters

|  | **≤ 10 Lap**<br>**Nephrectomies** | **> 10 Lap**<br>**Nephrectomies** | **p-value** |
| --- | --- | --- | --- |
|  | (n = 54) | (n = 27) |  |
| Respect of Tissue,<br>score [mean (SD)] | 3.76 (0.60) | 4.03 (0.42) | 0.073 |
| Time and Motion, score<br>[mean (SD)] | 3.55 (0.75) | 3.56 (0.62) | 0.765 |
| Instrument Handling,<br>score [mean (SD)] | 3.66 (0.69) | 3.77 (0.56) | 0.617 |
| Knowledge of<br>Instruments, score<br>[mean (SD)] | 4.02 (0.54) | 4.06 (0.58) | 0.491 |
| Flow of Operation,<br>score [mean (SD)] | 3.68 (0.63) | 3.81 (0.72) | 0.325 |
| Use of Assistant, score<br>[mean (SD)] | 3.69 (0.61) | 3.84 (0.62) | 0.246 |
| Procedure Knowledge,<br>score [mean (SD)] | 3.84 (0.68) | 3.97 (0.65) | 0.521 |
| Mean Overall Score<br>[mean (SD)] | 3.74 (0.56) | 3.86 (0.48) | 0.365 |

### Correlation between FLS Individual Task Scores and OSATS Mean Overall Scores

Mean overall OSATS scores correlated weakly with all three FLS tasks (Peg Transfer Timing: R = -0.331, R^2^ = 0.110; Precision Cutting Timing: R = -0.240, R^2^ = 0.058; Suturing with Intra-corporeal Knot Timing: R = -0.451, R^2^ = 0.203) (Figure 2, 3, 4). Although the correlations were statistically significant, this is likely due to the sizeable sample size that has allowed for identification of weak correlations in individual data.

**Figure 2:**
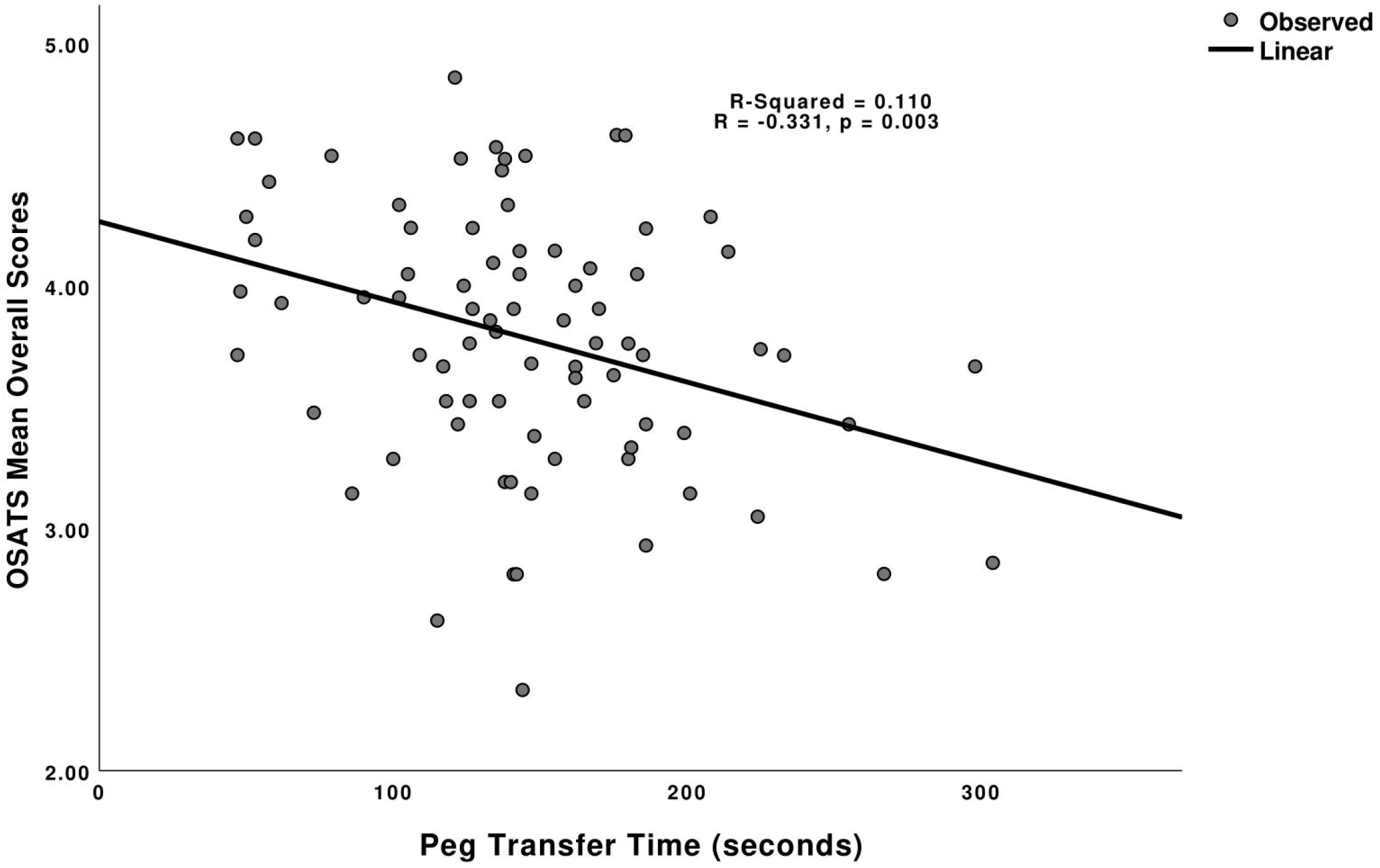
Correlation between Peg Transfer Time and OSATS Mean Overall Scores

**Figure 3:**
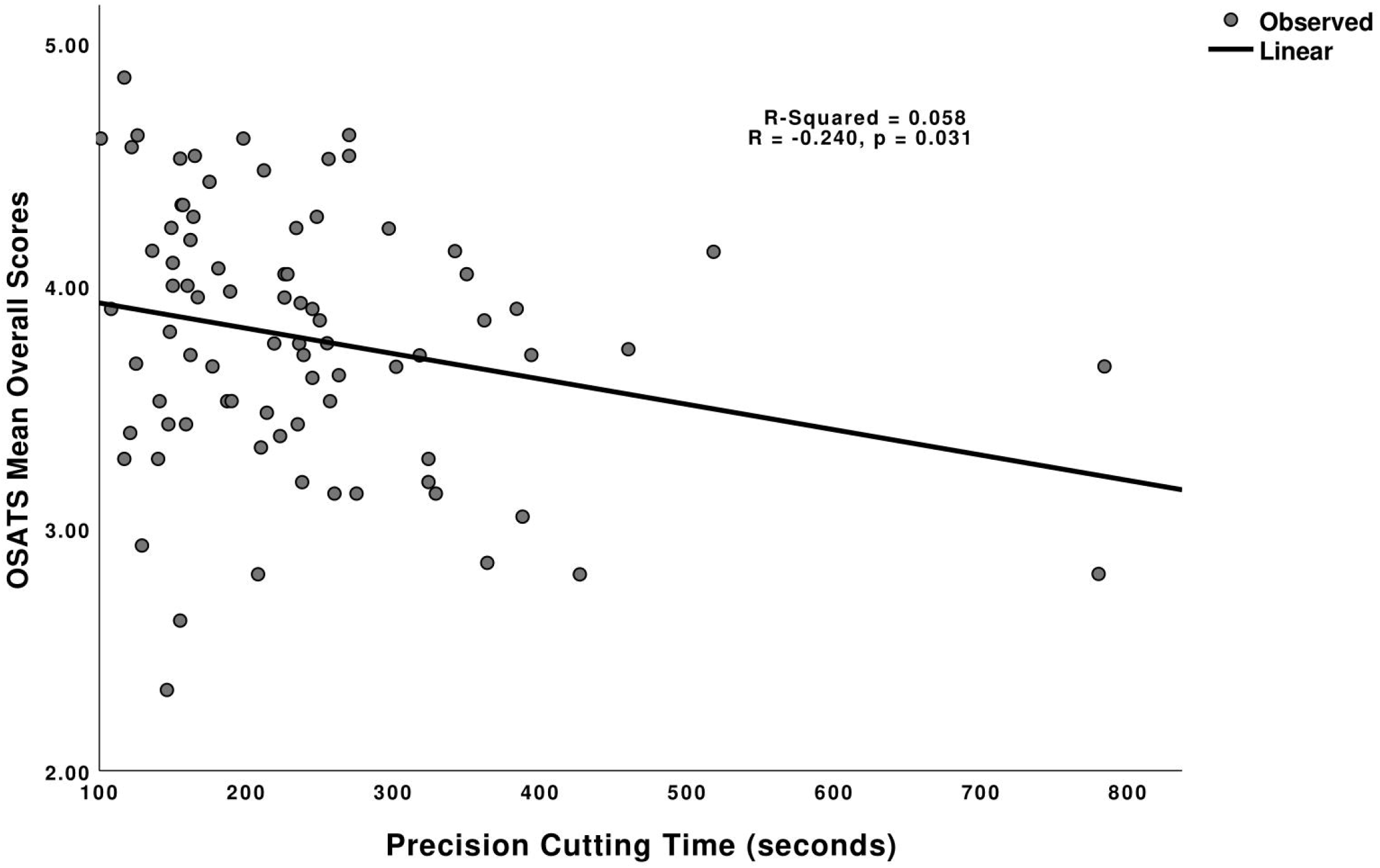
Correlation between Precision Cutting Time and OSATS Mean Overall Scores

**Figure 4:**
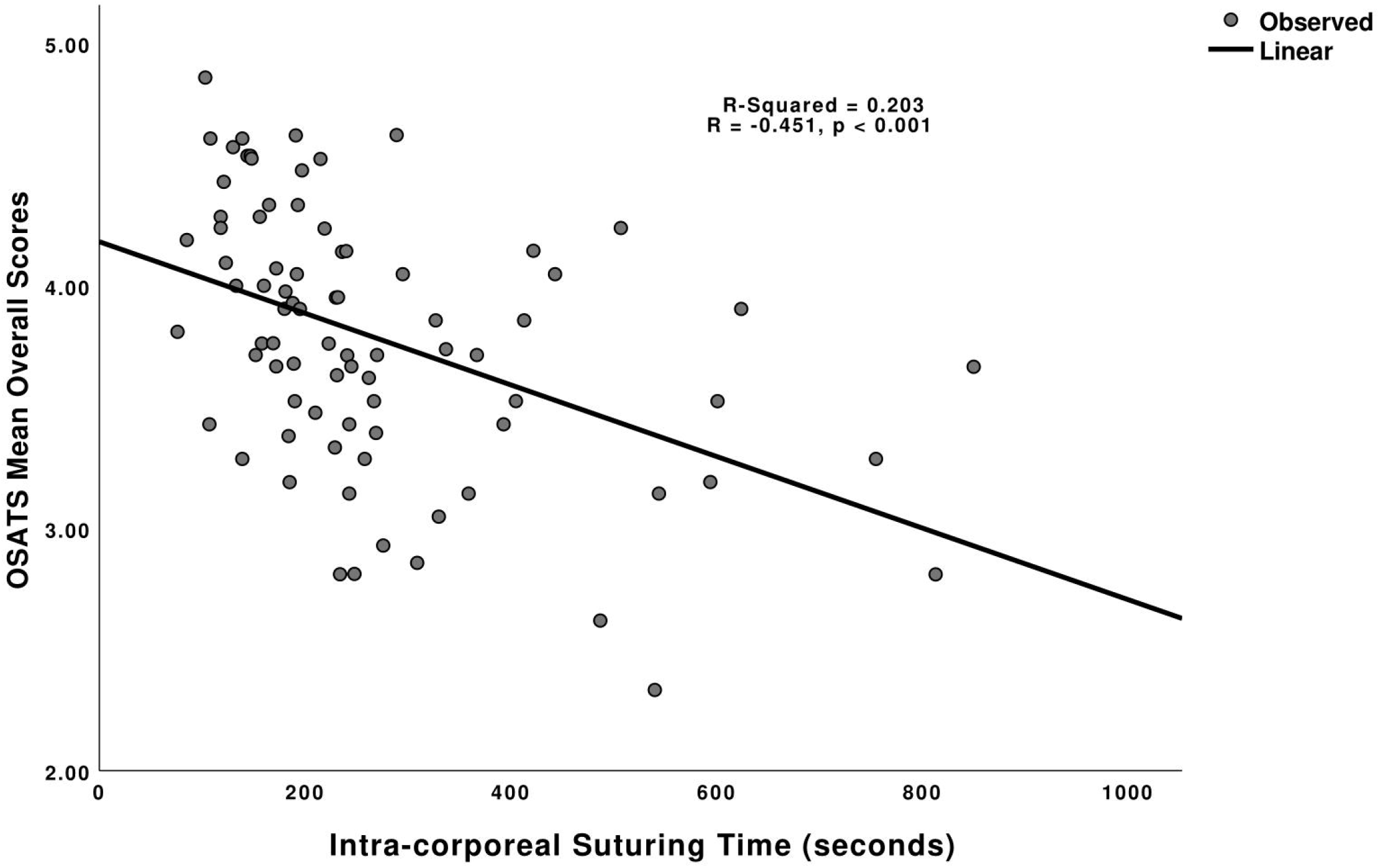
Correlation between Intra-corporeal Suturing Time and OSATS Mean Overall Scores

## DISCUSSION

Studies in current literature have extensively studied the relevance of FLS task simulations and OSATS global rating scales in assessing the proficiency of surgical trainees and professionals in laparoscopic skills. Not only have these tools been validated across various surgical specialties, but they have also been steadily refined and adapted to specific contexts and scenarios^9, 13, 19-21^. However, to the best of our knowledge, this is the first comparative study to evaluate the utility of FLS manual task parameters in predicting the performance of live porcine laparoscopic urological surgery as represented by OSATS scores on the global rating scale.

Our study demonstrated the absence of a strong correlation between FLS manual task timings and OSATS scores in live laparoscopic porcine surgery. In fact, this contrasts with published literature that has previously supported the predictive ability of FLS in assessing operative technical proficiency^12, 22, 23^. Although statistically insignificant, it is of worth noting that FLS task parameters and OSATS scores in our findings demonstrated an observable trend – participants with more experience generally accrued shorter timings in two out of three FLS tasks, and this trended with better OSATS scores in not only the individual components, but the mean overall score as well. After scrutiny of the data, it is possible that our enrolled target population did not notably differ in their laparoscopic experience, resulting in little variation across their scores even after stratification. For instance, there could have been a substantial number of participants that performed 11 nephrectomies to have been categorized within the group with more experience. Similarly, this may also be seen in the group with less experience, where a considerable number performed 10 nephrectomies, one less procedure away from being classified as having more experience. The converse is true for the studies referenced above, of which had a wide gulf of experience levels observed within their study population. Hence, the predictive validity reported in these cohorts are not surprising. It is also important to recognize that our study had no primer session for the delegates to familiarize themselves physically with the simulator task models prior to the course. A good proportion of the participants may have had their fledgling experience with the FLS simulators during the course itself, with minimal formal simulator training and exposure back in their home institution. Moreover, the live porcine surgery segment involved the assumption of the role of a primary surgeon between the 2 delegates working together, which may also turn out to be a novice opportunity for some in a setting of a laparoscopic operation. Such a proposition stems from the fact that a vast majority of our participants fell into the group with less experience, of which the lack of correlation may seek to suggest that the FLS task simulator models are not appropriate modes of evaluation in early surgeon trainees with minimum laparoscopic experience.

Our findings have also been supported by other studies^14, 24^ that discovered the underlying assumptions of how simulator-based training correlated with surgical skill competence. Anderson et al. concluded that OSATS scores did not correlate significantly with measured parameters during a simulation of intra-articular fracture reduction done amongst orthopedic residents, stating the need to customize specific assessment factors to accurately predict surgical outcomes. In this context, the nature of OSATS as an external evaluation could not comprehensively cover the procedure-specific metrics such as articular reduction quality and integrity of the mechanical fixation. Likewise, Steigerwald et al. demonstrated the lack of association between another laparoscopic skills global rating scale (Global Assessment of Laparoscopic Skills) and the FLS low-fidelity box trainer for porcine cholecystectomy, questioning the usability of such tools in the context of novice surgical trainees. The variances observed in literature thus seek to assert that simulation-based assessment may not necessarily always objectively signify surgical skill competence – many other confounding factors may potentially come into play and distort the seemingly direct association between these two variables.

## STRENGTHS AND LIMITATIONS

To the best of our knowledge, this is one of the only courses that has made simultaneous performance evaluation of participants in both technical and live operative aspects possible. All tasks are assessed in one sitting, minimizing any subjective performance disparities, and allowing for instant conveyance of technical skillsets into live surgery environment.

Although the Laparoscopic Upper Tract Surgery Course has been held consistently across three years, a larger sample size would undoubtedly aid in reducing any variability due to bias. Instead of comparing individual FLS task timings, an overall scoring metric may have allowed for a more direct comparison between the outcomes of interest. Lastly, owing to the unpredictable nature of surgeons’ schedules, the sessions organized from year to year may have seen varying attendances, which may have led to opportunistic sampling as a result. However, the probability of this occurring is rather slim due to the continued support and enrolment of the AUSTEG courses.

## CONCLUSION

Surgical simulator training has been consistently validated across different aspects of laparoscopic surgery. Likewise, FLS manual tasks have been evaluated alongside the use of these training models and regarded as highly reliable and accurate measures of laparoscopic skill in the operating room.

However, when evaluating the performance of live porcine laparoscopic urological surgery, FLS manual task parameters did not demonstrate strong correlations with overall OSATS scores. Despite being an established indicator of technical proficiency, this paper has demonstrated novel incongruencies between one’s performance in FLS technical laparoscopic skillsets and operative ability in the context of live porcine surgery. Discerningly, other variables are postulated to come into play – thorough knowledge of surgical anatomy, intricate operative techniques, and sufficient familiarity with relevant equipment are just some of many plausible reasons that determine the surgical prowess of an individual. Further studies exploring such contributory factors are highly anticipated.

Nevertheless, FLS remains to be a necessary adjunct to other objective assessment tools in evaluating operative aptitude but is admittedly not the be-all and end-all of assessing true competency in the operating theatre. Faculties should look towards incorporating a holistic, modular approach that aims to encompass both simulator-based and actual operative assessments to best delineate the laparoscopic skillset of a surgical candidate.

## Data Availability

All data produced in the present work are contained in the manuscript

## Acknowledgements

We confirm that our paper has not been previously published and is not currently under consideration for publication by other journals. We will not send our paper to another journal until a decision is made concerning publication.

All authors (Ho Yee Tiong, Wei Zheng So, Jeremy Yuen-Chun Teoh, Shuji Isotani, Gang Zhu, Teng Aik Ong, Eddie Shu-Yin Chan, Peggy Sau-Kwan Chu, Kittinut Kijvikai, Ming Liu, Bannakji Lojanapiwat, Michael Wong, Anthony Chi-Fai Ng) acknowledge that conflict of interests disclosures are complete for all named authors, to the best of their knowledge. All authors do not have any conflicts of interests to declare.

## REFERENCES

1. Fundamentals of Laparoscopic Surgery. https://www.flsprogram.org/.

2. Zheng B, Hur HC, Johnson S, Swanström LL. Validity of using Fundamentals of Laparoscopic Surgery (FLS) program to assess laparoscopic competence for gynecologists. Surg Endosc. 2010;24:152–160.

3. Seaman SJ, Jorgensen EM, Tramontano AC, et al. Use of Fundamentals of Laparoscopic Surgery Testing to Assess Gynecologic Surgeons: A Retrospective Cohort Study of 10-Years Experience. J Minim Invasive Gynecol. 2021;28:794–800.

4. Zendejas B, Ruparel RK, Cook DA. Validity evidence for the Fundamentals of Laparoscopic Surgery (FLS) program as an assessment tool: a systematic review. Surg Endosc. 2016;30:512–520.

5. Derossis AM, Fried GM, Sigman HH, Barkun JS, Meakins JL. Development of a model for training and evaluation of laparoscopic skills. The American journal of surgery. 1998;175:482–487.

6. Martin J, Regehr G, Reznick R, et al. Objective structured assessment of technical skill (OSATS) for surgical residents. Journal of British Surgery. 1997;84:273–278.

7. Hiemstra E, Kolkman W, Wolterbeek R, Trimbos B, Jansen FW. Value of an objective assessment tool in the operating room. Canadian Journal of Surgery. 2011;54:116.

8. Niitsu H, Hirabayashi N, Yoshimitsu M, et al. Using the Objective Structured Assessment of Technical Skills (OSATS) global rating scale to evaluate the skills of surgical trainees in the operating room. Surgery today. 2013;43:271–275.

9. Chang OH, King LP, Modest AM, Hur H-C. Developing an objective structured assessment of technical skills for laparoscopic suturing and intracorporeal knot tying. Journal of surgical education. 2016;73:258–263.

10. Fried GM, Feldman LS, Vassiliou MC, et al. Proving the value of simulation in laparoscopic surgery. Annals of surgery. 2004;240:518.

11. Datta V, Bann S, Beard J, Mandalia M, Darzi A. Comparison of bench test evaluations of surgical skill with live operating performance assessments. Journal of the American College of Surgeons. 2004;199:603–606.

12. McCluney A, Vassiliou M, Kaneva P, et al. FLS simulator performance predicts intraoperative laparoscopic skill. Surgical endoscopy. 2007;21:1991–1995.

13. Gumbs AA, Hogle NJ, Fowler DL. Evaluation of resident laparoscopic performance using global operative assessment of laparoscopic skills. Journal of the American College of Surgeons. 2007;204:308–313.

14. Anderson DD, Long S, Thomas GW, Putnam MD, Bechtold JE, Karam MD. Objective Structured Assessments of Technical Skills (OSATS) does not assess the quality of the surgical result effectively. Clinical Orthopaedics and Related Research®. 2016;474:874–881.

15. Hopmans CJ, den Hoed PT, van der Laan L, et al. Assessment of surgery residents’ operative skills in the operating theater using a modified Objective Structured Assessment of Technical Skills (OSATS): a prospective multicenter study. Surgery. 2014;156:1078–1088.

16. Teoh JY, Cho CL, Wei Y, et al. Surgical training for anatomical endoscopic enucleation of the prostate. Andrologia. 2020;52:e13708.

17. Teoh JY, Cho CL, Wei Y, et al. A newly developed porcine training model for transurethral piecemeal and en bloc resection of bladder tumour. World J Urol. 2019;37:1879–1887.

18. Rouach Y, Timsit MO, Delongchamps NB, et al. [Laparoscopic partial nephrectomy: urology resident learning curve on a porcine model]. Prog Urol. 2008;18:344–350.

19. Argun OB, Chrouser K, Chauhan S, et al. Multi-Institutional Validation of an OSATS for the Assessment of Cystoscopic and Ureteroscopic Skills. J Urol. 2015;194:1098–1105.

20. Kowalewski TM, Sweet R, Lendvay TS, et al. Validation of the AUA BLUS tasks. The Journal of urology. 2016;195:998–1005.

21. Halwani Y, Sachdeva A, Satterthwaite L, de Montbrun S. Development and evaluation of the general surgery objective structured assessment of technical skill (GOSATS). Journal of British Surgery. 2019;106:1617–1622.

22. Fried GM. FLS assessment of competency using simulated laparoscopic tasks. Journal of Gastrointestinal Surgery. 2008;12:210–212.

23. Dauster B, Steinberg AP, Vassiliou MC, et al. Validity of the MISTELS simulator for laparoscopy training in urology. Journal of endourology. 2005;19:541–545.

24. Steigerwald SN, Park J, Hardy KM, Gillman L, Vergis AS. The Fundamentals of Laparoscopic Surgery and LapVR evaluation metrics may not correlate with operative performance in a novice cohort. Med Educ Online. 2015;20:30024.

